# Intraspecies strain exclusion, antibiotic pretreatment, and donor selection control microbiota engraftment after fecal transplantation

**DOI:** 10.1101/2021.08.18.21262200

**Authors:** Daniel Podlesny, Marija Durdevic, Sudarshan Paramsothy, Nadeem O. Kaakoush, Christoph Högenauer, Gregor Gorkiewicz, Jens Walter, W. Florian Fricke

## Abstract

Fecal microbiota transplantation (FMT) is both a promising therapeutic approach to treat microbiota-associated pathologies and an experimental tool to establish a causal role of microbiome dysbiosis in human pathologies. Although clearly efficacious in resolving recurrent *Clostridioides difficile* infection (rCDI), the therapeutic value of FMT in other pathologies is not yet established, and our mechanistic and ecological understanding of how FMT alters the microbiome in patients is incomplete. Here, we assembled the most comprehensive FMT trial microbiota dataset to date, including new and previously generated fecal metagenomes from FMT trials in rCDI, inflammatory bowel disease (IBD), metabolic syndrome (MetS), drug-resistant pathogen colonization (MDR), and resistance to immune checkpoint inhibitor anti-tumor therapy (ICI). We characterized post-FMT microbiota assembly in the recipients by establishing the origin of the detected strains, and we identified the clinical and ecological factors that determine the engraftment of donor strains. Our findings showed little coexistence of donor and recipient strains and linked the magnitude of donor strain engraftment to dysbiosis of the recipient microbiome. Dysbiosis and strain engraftment were low in pathologies other than rCDI but could be enhanced through pretreatment with antibiotics and lavage. Using generalized linear mixed-effects models, we demonstrate that both ecological (low recipient and high donor ɑ-diversity and relative species abundance) and clinical (antibiotic pretreatment, bowel lavage, multiple rounds of FMT) variables are associated with increased donor microbiota engraftment, and that donor strain engraftment events are predictable for individual patients and strains. Overall donor strain engraftment was not linked to FMT outcome in IBD patients but was higher in ICI patients that responded to immunotherapy after FMT. Our findings provide an ecological framework for post-FMT microbiota assembly that can predict donor strain engraftment and determine its importance for clinical outcomes, informing more targeted and personalized approaches to increase the therapeutic benefits of FMTs.

## INTRODUCTION

Fecal microbiota transplantation (FMT) represents the clinical attempt to treat diseases that are associated with a disturbed gut microbial ecosystem, often referred to as “dysbiosis” (Hooks and O’Malley 2017), by infusing healthy donor feces into the intestinal tract of a patient (D’Haens and Jobin 2019). FMT has been validated in randomized controlled trials (RCTs) as a treatment for recurrent *Clostridioides difficile* infection (rCDI) with a success rate of ∼90%, surpassing that of conventional antibiotic treatment (Kao et al. 2017; van Nood et al. 2013). FMT efficacies in inducing remission in IBD patients with ulcerative colitis have generally been lower (24–32% versus 5–9% for placebo), but surpassed those reported in phase III clinical trials for several biological agents (golimumab and vedolizumab) (Danne, Rolhion, and Sokol 2021). Two recent trials suggest a potential role for FMT in cancer therapy, demonstrating re-induction of a response to anti-PD-1 immunotherapy in ∼30% of immune checkpoint inhibitor (ICI)-refractory melanoma patients (n=25) after FMT from patients that had previously responded to ICI (Davar et al. 2021; Baruch et al. 2021). Numerous RCTs are under way to test FMT as a treatment for other microbiota-associated infectious, inflammatory, and metabolic diseases. From a microbiome research perspective, FMT represents an intriguing experimental model to establish causality in reported human microbiota/disease associations, which are currently mostly studied using controversial germ-free mice experiments (Walter et al. 2020). Yet, the mechanism of action of FMTs and their specific and long-term effects on the recipient microbiota remain poorly understood (Khoruts, Staley, and Sadowsky 2021; Haifer et al. 2021).

The population dynamics of post-FMT microbiota organization in patients from different medical backgrounds have not been extensively studied, and the clinical, host, and ecological factors that govern microbiome assembly at the level of individual patients and individual microbes are insufficiently understood (Danne, Rolhion, and Sokol 2021). The vast majority of studies were based on 16S rRNA analysis, which lacks strain-level resolution and is unable to specifically track recipient and donor-derived microbiota contributions. Given that competitive interactions among microbiome members are hypothesized to be highest among closely related taxa, including different strains of the same species (Walter, Maldonado-Gómez, and Martínez 2018), strain-level resolution is a prerequisite to understanding the ecological processes of post-FMT microbiome assembly. Two previous studies applied strain-level profiling methods to characterize microbiota engraftment in FMT-treated metabolic syndrome (n=5; (Li et al. 2016)) and rCDI (n=18; (Smillie et al. 2018)) patients. Li et al. found a “durable coexistence of donor and recipient strains” within the same species in metabolic syndrome patients after FMT, and a higher chance for donor strain engraftment, if another strain from the same species was already present in the recipient before FMT (Li et al. 2016). In rCDI patients, Smillie et al. found less evidence for substantial and lasting recipient and donor strain coexistence, which was only observed in some patients, but reported engraftment of multi-strain donor species populations in an “all-or-nothing manner” (Smillie et al. 2018). These findings appear inconsistent with the competitive intraspecies exclusion that would be expected for closely related strains with overlapping resource niches based on coexistence theory (Letten, Hall, and Levine 2021). Competitive intraspecies strain exclusion as a governing principle for gut microbial ecosystems would be further supported by studies that found most species of the human fecal microbiota to be dominated by a single strain (Truong et al. 2017) and orally supplemented probiotic strains to only engraft if a closely related strain was absent in the resident microbiome (Maldonado-Gómez et al. 2016). Coexistence theory has, to our knowledge, not been applied to understand the effects of FMTs, and there is an incomplete understanding of the strain-level dynamics of post-FMT microbiota assembly, including the factors that control donor strain engraftment, patient strain persistence, and recipient/donor strain competition. An improved knowledge of these mechanisms will be needed to predict and control FMT outcomes, determine the relationship of post-FMT microbiota assembly and clinical response, and define the opportunities and limitations of FMT-based microbiota therapies.

Although FMT represents an ecosystem restoration approach in many aspects, ecological framework theories have only recently been applied to understand and predict FMT outcomes (Xiao et al. 2020). The healthy adult gut microbiota exhibits colonization resistance to invading microbes that compete for ecological niches that are shared with resident microbes (Lawley and Walker 2013; Stecher and Hardt 2008). A dysbiotic microbiota (e.g. after antibiotic treatment) provides reduced colonization resistance to *C. difficile* infection (Battaglioli et al. 2018) and is unable to recover in rCDI patients due to repeated rounds of unsuccessful antibiotic treatments (Song et al. 2013), allowing for the establishment of a transplanted donor microbiota in rCDI patients after FMT (Smillie et al. 2018; Podlesny and Fricke 2020)(Podlesny et al., submitted). To what extent a donor microbiota can establish in patients with pathologies that do not present with a severely disrupted gut microbiome remains unclear. Furthermore, the ecological framework that controls donor microbiota engraftment and determines the impact of clinical modalities on post-FMT microbiota assembly and FMT outcome has not been resolved.

In order to identify the critical microbiome and clinical determinants of post-FMT microbiota assembly, we sought to systematically identify and compare donor-derived microbiota fractions in FMT-treated patients in relation to variable disease and treatment backgrounds. We found antibiotic treatment before FMT, as part of the patient history or pretreatment for FMT, and resulting dysbiosis signatures to have the strongest positive impact on donor microbiota engraftment. We further developed models to predict donor microbiota engraftment in individual patients and for individual strains, which align with ecological theory, and demonstrated a major potential for personalized FMT outcome optimization based on both simulated strain-supplemented donor samples and recipient/donor pairs from our meta-cohort. Finally, we show evidence that increased contributions of donor-derived strains to the post-FMT patient microbiota are associated with the induction of a response to immunotherapy in ICI-refractory melanoma patients but not with remission in IBD patients.

## RESULTS

### Fecal microbiota changes and dysbiosis signatures in a human meta-cohort of FMT-treated patients with different medical conditions

To compare the effect of FMT on fecal microbiota composition in patients with different medical conditions, a comprehensive meta-cohort was assembled of newly generated and available metagenomic shotgun sequence data from FMT-treated patients and donors, including 1232 samples from 245 cases (Table S1). This FMT meta-cohort (Fig. 1A) includes patients treated for recurrent *C. difficile* infection (rCDI, 3 studies), the inflammatory bowel diseases (IBD) ulcerative colitis (2 studies) and Crohn’s disease (1 study), metabolic syndrome, type 2 diabetes mellitus or obesity (MetS, 3 studies), drug-resistant *Enterobacteriaceae* carriage (MDR, 2 studies), or immune checkpoint inhibitor-refractory melanoma (ICI; 2 studies). Post-FMT samples were relatively evenly distributed between patient groups over the first four months after FMT (Fig. S1). For comparison, a control cohort of healthy individuals from several unrelated fecal microbiome studies was included (See Methods for detailed description). There were substantial differences between FMT study protocols, including the use of bowel lavage and antibiotic treatment (ABx^+/-^), the FMT application type (by nasoduodenal or colonic endoscopy, enema or capsule), as well as the number of FMTs received (which varied between a single and up to 41 applications). All rCDI patients were treated with antibiotics as part of their therapeutic regimens, while each of the ICI, IBD, and MDR groups included at least one study where antibiotics were used as a bowel cleansing strategy to deplete the resident microbiota and prepare patients for donor microbiota engraftment (Fig. 1A). This meta-cohort reflects the current state and heterogeneity of patient populations and disease backgrounds that have been experimentally treated with FMT.

**Fig. 1.**
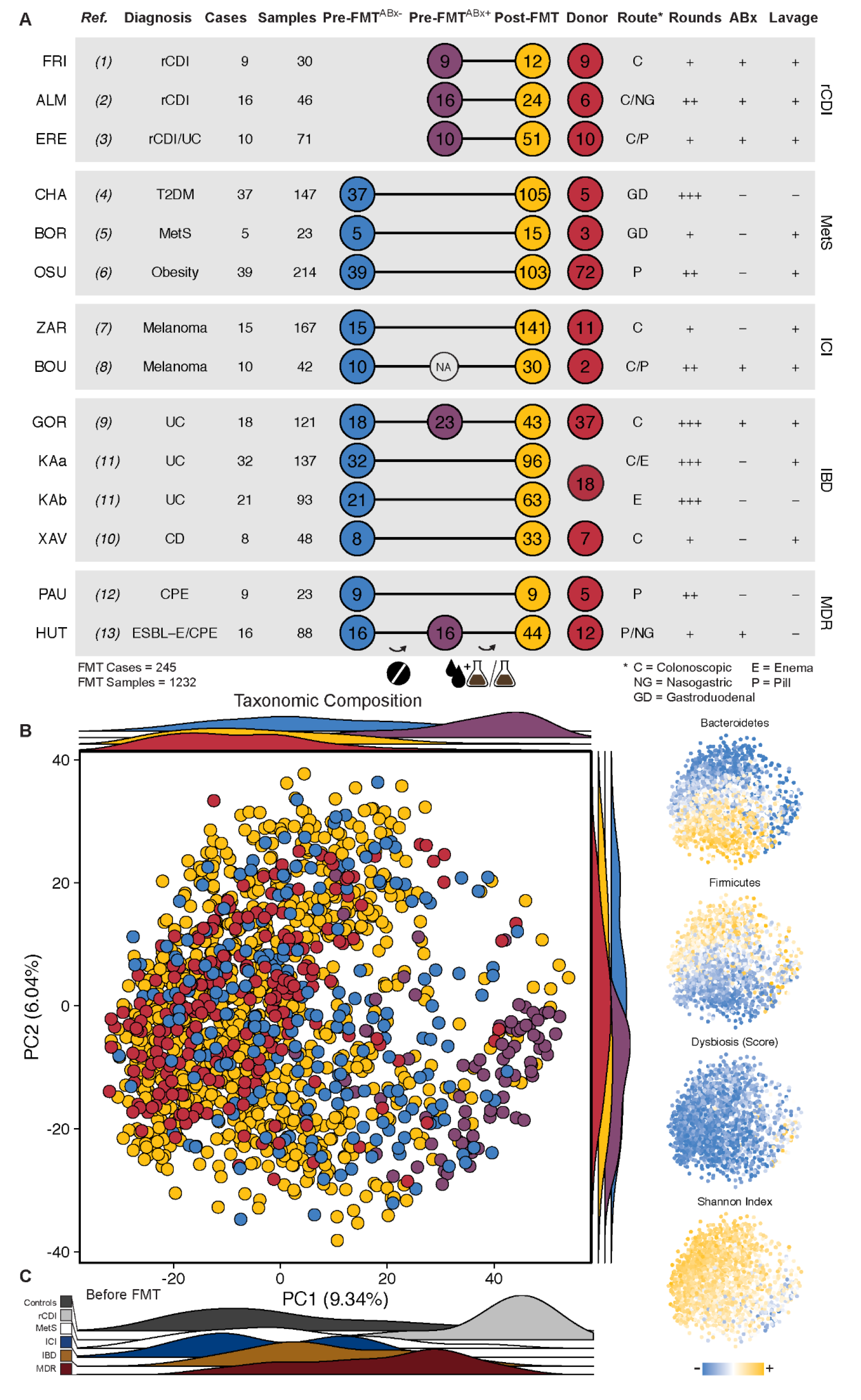
Overview and taxonomic microbiota composition of the FMT meta-cohort. **A**) Overview of treatment modalities, number and distribution of cases and samples for thirteen studies from five conditions included in the FMT meta-cohort. **B**) Taxonomic microbiota compositions based on principal component analysis (PCA) of centered log-ratio-transformed relative species abundance. Samples in the main plot are categorized and color-coded as pre-FMT^ABx-^ (blue), antibiotically pretreated pre-FMT^ABx+^ (purple) and post-FMT (yellow) patient and donor (red) samples. Ridgeline density plots show sample distributions along the two first principal components based on the same categories. In the small plots (right side) samples from the same PCA are color-coded based on (from top to bottom) scaled cumulative relative abundances of *Bacteroidetes*, *Firmicutes*, dysbiosis score (Gevers et al. 2014), and alpha-diversity (Shannon index). **C**) Ridgeline density plots showing only pre-FMT (untreated and antibiotically pretreated) patient samples, colored by disease category.

We applied principal component analysis of a beta-diversity distance computed with centered log ratio-transformed relative species abundances to characterize the variation in fecal microbiota composition over the entire cohort (Fig. 1B). Taxonomic compositional microbiota differences in the data were best explained by principal component (PC) 1. The separation of samples along PC1 correlated with alpha-diversity (Shannon diversity) and a dysbiosis score based on relative species abundances (Gevers et al. 2014), whereas PC2 primarily reflected shifts in the *Bacteroidetes*/*Firmicutes* ratio between samples (Fig. 1B, inlet). Before FMT, rCDI patient samples largely separated from other pre-FMT patient samples (Fig. 1C), whereas the pre-FMT microbiota of other patients not treated with antibiotics was clustered with that of donors and healthy controls (Fig. 1B, C), showing that these medical conditions were not associated with a pronounced dysbiosis. In contrast, antibiotic treatment before FMT placed IBD and MDR patient samples in the dysbiotic range of rCDI patient samples (Fig. 1B). Given that rCDI is characterized by repeated antibiotic treatment attempts to eradicate *C. difficile* infection (Leffler and Lamont 2015), our analysis suggests that antibiotics are the main drivers of the compositional microbiota variations shown on PC1 and result in similar alterations to the microbiota, independently of the patient medical condition.

To better define dysbiosis signatures in our FMT meta-cohort, all patient and donor samples were compared based on taxonomic and functional microbiota parameters and patient samples further divided into antibiotically treated and untreated samples, based on study protocols and available sequence data (Fig. 2). As suggested by the PCA, rCDI^ABx+^ patient samples consistently exhibited decreased alpha-diversity, increased taxonomic distance relative to healthy controls, and elevated dysbiosis scores, whereas similar dysbiosis signatures were not observed in patients from all ICI^ABx-^, IBD^ABx-^ and, with the exception of one study, MetS^ABx-^ cohorts. However, IBD^ABx+^ patients adopted a dysbiosis state comparable to rCDI patients after broad-spectrum (vancomycin, paromomycin and nystatin) antibiotic pretreatment (Fig. 2, GOR; (Kump et al. 2018)). A similar trend was observed in MDR patients, although these patients were difficult to classify as they included antibiotically treated patients following 48 hours of antibiotic discontinuation ((Bar-Yoseph et al. 2020; Leo et al. 2020)) or received antibiotics specific for only Gram-negative bacteria (HUT, colistin and neomycin; (Huttner et al. 2019)). Functional dysbiosis was characterized by increased cumulative relative abundances of oral and oxygen-tolerant species in rCDI^ABx+^, IBD^ABx+^, and MDR^ABx+^ patient samples (Fig. 2). Dysbiosis signatures were resolved or at least improved in rCDI patients and absent from all other patient populations after FMT. Thus, microbiota-associated diseases besides rCDI that have been experimentally treated with FMT are characterized by low levels of dysbiosis, but rCDI-like dysbiosis is induced after antibiotic treatment, with likely consequences for the patient’s colonization resistance to invading microbes (S. Kim, Covington, and Pamer 2017).

**Fig. 2.**
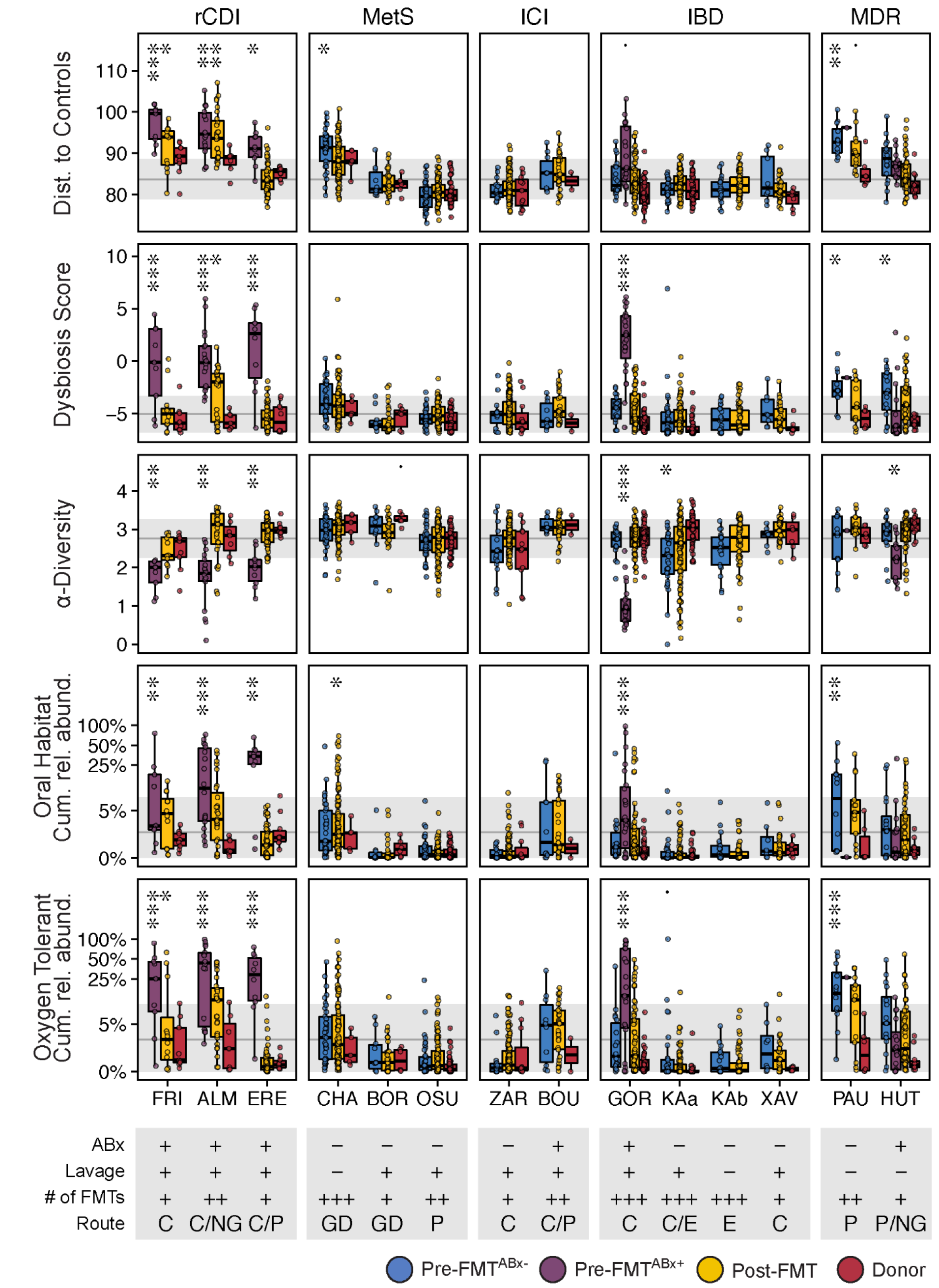
Taxonomic and functional microbiota comparison of FMT recipients, with or without antibiotic treatment, post-FMT patients, and donors in the different studies. Generalized linear mixed-effects models (GLMM, see Methods) highlight differences in taxonomic and functional microbiota metrics between Pre-FMT^ABx-^ (blue), pretreated Pre-FMT^ABx+^ (purple), Post-FMT (yellow), and donor (red) samples relative to a reference cohort of 739 healthy adults (grey line and area denote mean ± s.d., respectively), based on the average distance to healthy control samples (β-diversity, Aitchison distance), the dysbiosis score by (Gevers et al. 2014), α-Diversity (Shannon index), and the cumulative relative abundance of oxygen-tolerant or oral bacterial species. Metadata abbreviations indicate pretreatment with ABx and lavage (+/-); single (+), two (++), or multiple (+++) FMTs; colonoscopic (C), nasogastric (NG), or gastroduodenal (GD) FMT route, enema (E), and pill/capsule (P) administration. Significant differences of the different sample types from healthy controls were determined for each metric and study separately with generalized linear mixed-effects models. Asterisks denote significance thresholds: ̇p≤.1, *p≤.01, **p≤.001, ***p≤.0001.

### Strain tracking resolves variable and patient pretreatment-dependent donor strain contributions to the post-FMT microbiota

The pretreatment microbiome in recipients of FMTs was dominated by species detected in both donors and recipients that accounted for >75% cumulative relative abundance (Fig. 3A, inlet). Assigning specific members of the post-FMT patient microbiota to recipient and donor sources therefore requires taxonomic profiling below the species level. Contributions of donor and recipient strains to the post-FMT patient microbiota were identified with the SameStr tool from our group (Podlesny and Fricke 2020)(Podlesny et al., submitted), which has recently been validated in a neonatal microbiota meta-analysis (Podlesny and Fricke 2021). SameStr allows for the reliable detection of recipient-derived strains as shared strains between pre- and post-FMT patient metagenomes or donor-derived strains as shared strains between post-FMT patient and donor metagenomes (Fig. 3A). Importantly, SameStr applies a more conservative threshold for strain calls than related tools (Podlesny and Fricke 2020)(Podlesny et al., submitted). This allowed us to unambiguously assign post-FMT patient strains to recipient or donor sources, whereas less conservative strain definitions can include more broadly present subspecies lineages that are also shared between unrelated individuals or samples (Podlesny and Fricke 2020)(Podlesny et al., submitted). To validate the specificity of SameStr in our FMT meta-cohort, we determined the “false-positive” shared strain detection rate in 2606 sample pairs from distinct individuals that would not be expected to share or have exchanged microbial strains (both donors and post-FMT patients) (Fig. 3B). This analysis identified only 0.4 ± 1.1 shared strains in unrelated sample pairs, attesting to the high specificity of this method for the identification of case-specific or unique shared strains, which is needed to infer donor strain engraftment in post-FMT patients.

**Fig. 3.**
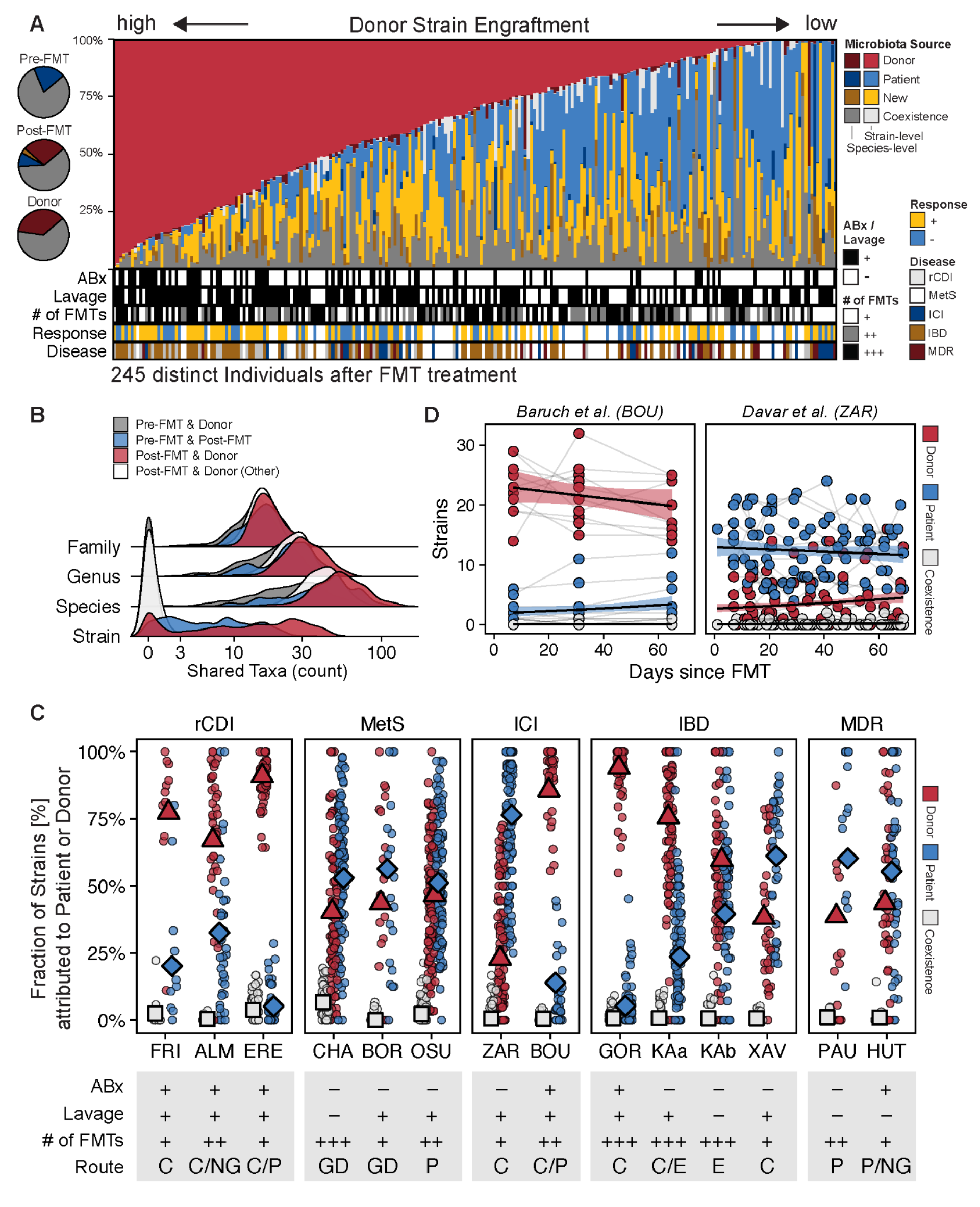
Strain profiling in FMT recipients showed substantial variation in donor strain engraftment between studies that is linked to FMT treatment modalities. **A**) Post-FMT microbiota relative abundance fractions contributed from donor (red), patient (blue), new (yellow), or coexisting (grey) strains, as detected in the last available post-FMT sample per patient. Darker colors refer to species-level relative abundance fractions if strains could not be resolved. *Left*: Circle chart showing average cumulative relative abundances of shared species in pre- and post-FMT patient and donor samples (grey) and between pre- and post-FMT patient (blue) and post-FMT patient and donor (red) samples. **B**) Validation of SameStr’s specificity to infer donor strain engraftment from shared strain detection. Very few shared strains were identified between pre-FMT patient and donor (grey) and between unrelated post-FMT patient and donor (white) sample pairs (considered false-positive shared strain calls), whereas strain sharing is frequent between pre- and post-FMT patient (blue) and between post-FMT patient and corresponding donor (red) sample pairs. The microbiota compositions of all sample pairs overlapped widely at higher taxonomic levels (family, genus, species). **C**) Comparison of donor-derived (red), recipient-derived (blue), and coexisting (grey) strain fractions in post-FMT patient samples (of the sum of donor and recipient-derived strains) between disease groups and individual studies from our meta-cohort. Symbols denote the mean value of the latest available post-FMT sample per patient and across all cases of a study. Study metadata are shown as follows: Antibiotic (ABx) and bowel lavage patient pretreatment: Yes (+), No (-); Number of FMTs: single (+), two (++), or multiple (+++) rounds; FMT application: by colon (C), nasogastric (NG), or gastroduodenal (GD) endoscopy, enema (E), or pill/capsule (P). **D**) Longitudinal comparison of donor-derived (red), patient-derived (blue), and coexisting (white) strain fractions in post-FMT samples from antibioticially pretreated (BOU) and non-pretreated (ZAR) ICI-refractory melanoma patients.

Application of SameStr to determine the origin of strains in recipients after FMT revealed that coexistence between donor- and recipient-derived strains, although detectable in 27.3% of post-FMT patients, only involved 0.6% of all species detected in patients post-FMT and accounted for only 2.2 ± 5.6% cumulative relative abundance. Importantly, recipient and donor strain coexistence was equally rare across all compared studies and disease backgrounds (Fig. 3C), including MetS patients for which extensive coexistence had previously been reported based on a different strain analysis method (Li et al. 2016). As the emergence of strain coexistence after FMT is dependent on donors and recipients carrying the same species before FMT, we specifically studied these cases of recipient and donor strain competition in our meta-cohort. Of 7641 cases where the same species was detected in donors and patients before FMT, donor and recipient strain coexistence was identified in only 127 cases (1.7 %), whereas either the donor or recipient strain was exclusively detected in 953 (12.5 %) and 1334 cases (17.5 %), respectively. In 2770 cases (36.3 %), post-FMT patients carried a new, previously undetected strain that could not be tracked back to the recipient or donor. In the remaining 2457 cases (32.2%), the species was no longer detected (1304 cases, 17.1%) or could not be analyzed at strain-level resolution (1153 cases, 15.1%). In summary, coexistence of recipient and donor strains after FMT might be less common than previously suggested (Li et al. 2016), and the presence of distinct strains from the same species in recipients and donors before FMT appears to predominantly lead to strain competition and mutual exclusion during post-FMT microbiota assembly.

Most strains detected in patients post-FMT could be assigned to either donor or recipient sources or were identified as new strains. Overall, donor-derived strains accounted for the largest fraction of the post-FMT patient microbiota (18.5 ± 26.4%), followed by recipient-derived (12.5 ± 20.3%), but donor and recipient-derived strain and relative abundance fractions varied substantially between cases and studies (Fig. 3A, C). While donors consistently contributed larger strain fractions to post-FMT rCDI patients (76.5 ± 27.1% of detected donor and recipient-derived strains), the post-FMT microbiota of non-antibiotically pretreated MetS^ABx-^, IBD^ABx-^ and ICI^ABx-^ patients was dominated by recipient strains. Interestingly, donor-derived strains became dominant after FMT in IBD and ICI in the two studies that applied antibiotic treatment regimens to prepare patients for FMT (Fig. 3C). The enhancing effects of antibiotic pretreatment on donor strain engraftment were particularly striking in ICI patients (Fig. 3D), where donor-derived strains accounted for 85.6 ± 15.7% of all strain observations in ICI^ABx+^ patients (Baruch et al. 2021) compared to only 23.0 ± 23.9% in ICI^ABx-^ patients (Davar et al. 2021). Donor and recipient-derived, as well as coexisting, strain contributions to the post-FMT patient microbiota remained remarkably stable in both ICI studies for at least 60 days after FMT (Fig. 3D).

Different antibiotic pretreatments had variable effects on post-FMT donor strain engraftment, which were linked to patient dysbiosis before FMT: MDR^ABx+^ patients treated with antibiotics specific for Gram-negative bacteria (colistin and neomycin) acquired smaller proportions of donor-derived strains (HUT, 43.7 ± 32.9%) than IBD^ABx+^ patients treated with an antibiotic cocktail against Gram-positive bacteria, parasites and fungi (vancomycin, paromomycin, and nystatin) (GOR, 94.1 ± 8.8%) or ICI^ABx+^ patients treated with vancomycin and neomycin (BOU, 85.6 ± 15.7%) (Fig. 3C). Interestingly, increased donor strain engraftment in the IBD^ABx+^ compared to the MDR^ABx+^ patients was also associated with enhanced pretreatment-induced dysbiosis before FMT (see GOR and HUT in Fig. 2), although the lack of additional pre-FMT^ABx+^ sequence data in the meta-cohort prevented a more comprehensive analysis.

Other forms of patient pretreatment, as well as the route of FMT application, likely also affected post-FMT microbiota assembly. Donor-derived strain contributions were larger in the KAa subgroup of IBD patients from the study by (Paramsothy et al. 2019) that received bowel preparation by lavage, followed by colonoscopic FMT and multiple enemas (75.6 ± 21.3%), compared to the second KAb subgroup that was treated with multiple enemas alone, without lavage and colonoscopic FMT (59.7 ± 27.5%) (Fig. 3C). Disruption of the patient microbiota in preparation for FMT, in particular with antibiotic therapy against Gram-positive bacteria, therefore appears to induce dysbiosis and enhance donor strain engraftment after FMT, including in different disease contexts.

### Microbiota and clinical determinants of donor strain engraftment after FMT in individual patients

In order to gain a conceptual understanding of the factors that determine donor microbiota engraftment after FMT in individual patients, we wanted to determine the effects of both ecological microbiota and clinical FMT parameters on post-FMT patient microbiota assembly. As ecological parameters, recipient and donor microbiota (ɑ/β) diversity, and as clinical parameters, patient pretreatment (ABx, lavage) and FMT modalities (no. of FMTs) were used as input variables for a generalized linear mixed model (GLMM) to estimate donor-derived strain fractions after FMT per patient (Fig. 4A, Table S8).

**Fig. 4.**
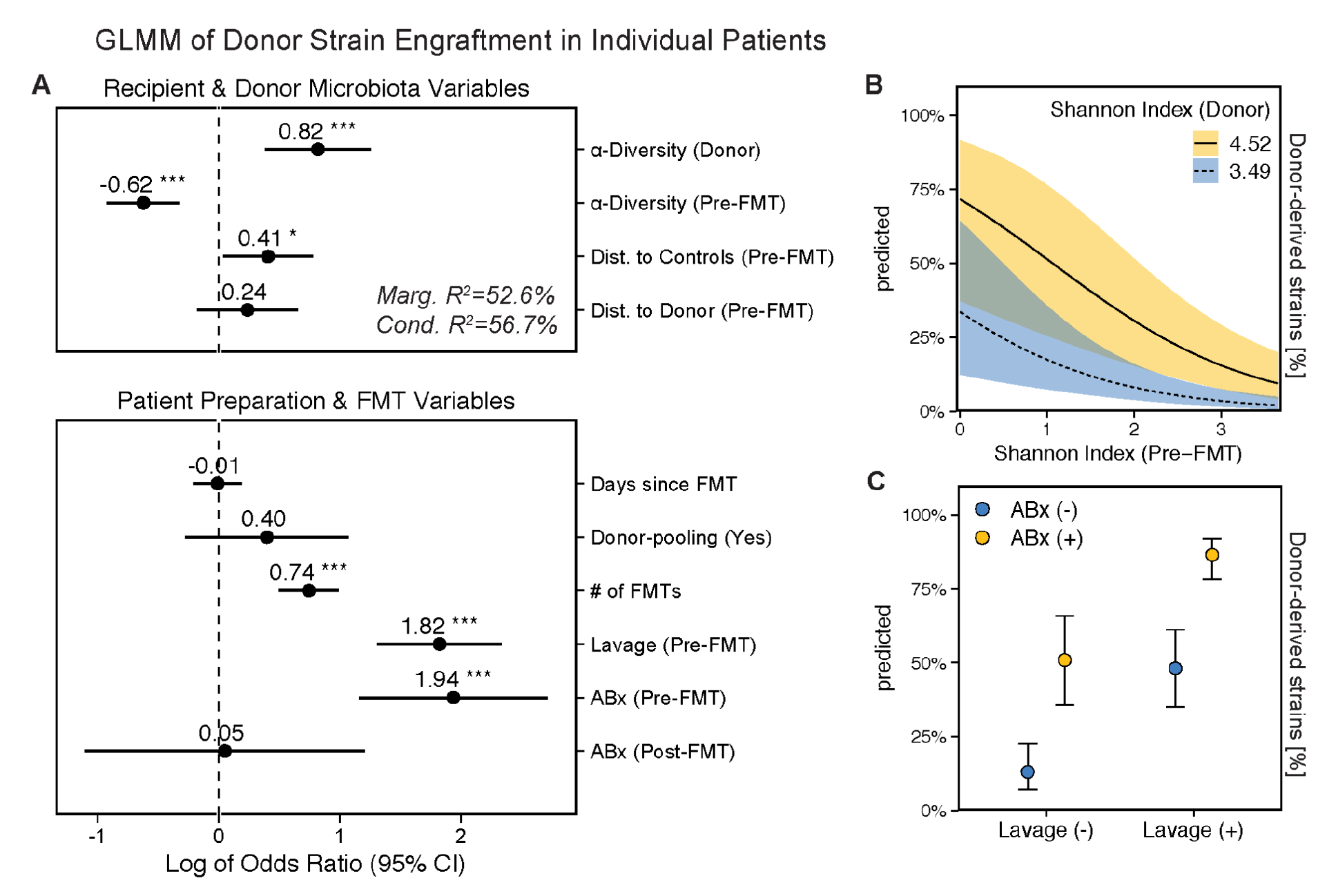
For individual patients, donor microbiota engraftment after FMT is dependent on patient and donor microbiota characteristics and clinical modalities of the FMT treatment. **A**) Forest plot showing the relevance of microbiota and clinical parameters for donor-derived strain fractions in post-FMT patients (see Fig. 3C) in the FMT meta-cohort, as determined with a generalized linear mixed model (see Methods). **B**) Simulations with this model to determine the marginal effects of α-diversity on donor strain engraftment, i.e. using real values in combination with the minimum or maximum Shannon index detected in any donor in the FMT cohort (min/max within 95% confidence intervals), indicate a disproportionate impact of high-α-diversity donors on low-α-diversity FMT recipients. **C**) Similar simulations predict independent marginal effects of ABx and lavage pretreatment on donor strain engraftment. Shaded areas and bars denote the confidence interval. Asterisks denote significance thresholds: *p≤.01, **p≤.001, ***p≤.0001.

Of the microbiota variables, donor ɑ-diversity (Shannon index) had the strongest positive predicted effect (OR=.82, p<.001), whereas recipient ɑ-diversity had a negative predicted effect (OR=-.62, p<.001) on donor strain engraftment. To estimate the marginal effects of recipient and donor ɑ-diversity alone, FMT outcomes were simulated for all real recipient/donor cases but with different donor ɑ-diversity values. When the actual donor ɑ-diversity was replaced alternatively with the highest and lowest Shannon index detected in any donor from the meta-cohort, simulations indicated that a disproportionately large fraction of strains from high-diversity donors would engraft in low-diversity patients after FMT (Fig. 4B). Compositional divergence (β-diversity, Aitchison distance) of the recipient relative to the control microbiota (healthy reference cohort) - a microbiota marker for dysbiosis (Fig. 2) - was positively correlated with donor microbiota engraftment (OR=.41, p<.05). Compositional distance between recipient and donor microbiota, however, was not predicted to significantly affect engraftment (OR=.24, p=ns), suggesting that donor strain engraftment is more dependent on recipient dysbiosis and donor and recipient α-diversity than specific donor microbiota compositions.

Of the clinical variables, antibiotic pretreatment (OR=1.94, p<.001), bowel lavage (OR=1.82, p<.001), and multiple FMT applications (OR=.74, p<.001) were all predicted to increase donor strain engraftment based on our GLMM (Fig. 4A). Time since FMT had no effect on engraftment (OR=-.01, p=.914), suggesting temporally stable donor contributions to post-FMT microbiota assembly. Donor sample pooling was also not predicted to increased donor strain engraftment (OR=.40, p=.248), which is surprising given the positive association of engraftment with donor ɑ-diversity and could indicate that a mixture of multiple lower-microbiota diversity samples is not functionally equivalent to a single high-microbiota diversity sample for FMT. FMT outcome predictions with the GLMM using artificial ABx treatment and lavage parameters in the context of real values for all other parameters indicate that both pretreatment measures independently increase the rate of donor strain engraftment after FMT (Fig. 4C).

Thus, post-FMT microbiota assembly appears to be driven both by the recipient (ɑ/β-diversity) and donor (ɑ-diversity) microbiota, as well as the FMT procedure, i.e. resident microbiota depletion before FMT (by ABx treatment and lavage) and repeated exposure of the patient to the donor microbiota (multiple FMTs).

### Impact of recipient/donor matching on the engraftment probabilities of individual strains

The previous model estimated FMT outcomes for individual patients based on donor-derived post-FMT microbiota fractions. Next, we sought to model and compare the engraftment probabilities for individual strains from different microbial taxa. Our goal was to develop a framework that could outline the taxonomic boundaries for donor strain engraftment, i.e. whether strains from specific taxa would be more or less likely to engraft after FMT and which taxonomic and functional properties, as well as microbiome-dependent factors and clinical variables, would influence their engraftment probabilities. We used a GLMM to determine the importance of the same set of microbiota and clinical variables, as described above, for the engraftment of individual strains, but in combination with microbial species properties. These microbial properties (relative abundance, Gram stain, oral habitat, spore formation, and oxygen tolerance) were aggregated at the genus level to provide us with a more robust statistical foundation and a single, generalizable model for the entire meta-cohort.

Estimated donor strain engraftment probabilities were generally high for members of the phylum *Bacteroidetes*, low for *Proteobacteria,* and mixed for *Firmicutes* and *Actinobacteria* (Fig. 5A). On the genus level, the median estimated engraftment probability was 0.92%, with *Megamonas* (21.7%), *Desulfovibrio* (16.8%), and *Paraprevotella* (13.7%) representing the most and *Klebsiella*, *Veillonella*, and *Haemophilus* (<2e-07%) the least likely engrafted genera (Fig. 5B). Oral habitat (OR=-.81, p<.001), spore formation (OR=-.12, p<.001), and oxygen tolerance (OR=-.11, p<.05) were all negatively correlated with engraftment probability (Fig. 5C, Table S9). Across all detected microbial genera, donor strains were more likely to engraft if the corresponding species had a higher relative abundance in the donor (OR=1.51; p<.001), but less likely if the corresponding species had a higher relative abundance in the recipient (OR=-.06, p<.05) or if the species relative abundance was higher in the recipient relative to the donor (‘Interaction’, OR=-.07, p<.001). In conclusion, the engraftment of specific donor strains appears to be less dependent on microbiome (ɑ/β-diversity) and clinical (ABx, lavage) parameters, but specific properties of the associated species, such as (oral, spore-forming, oxygen-tolerant) lifestyle and (recipient and donor) relative abundance.

**Fig. 5.**
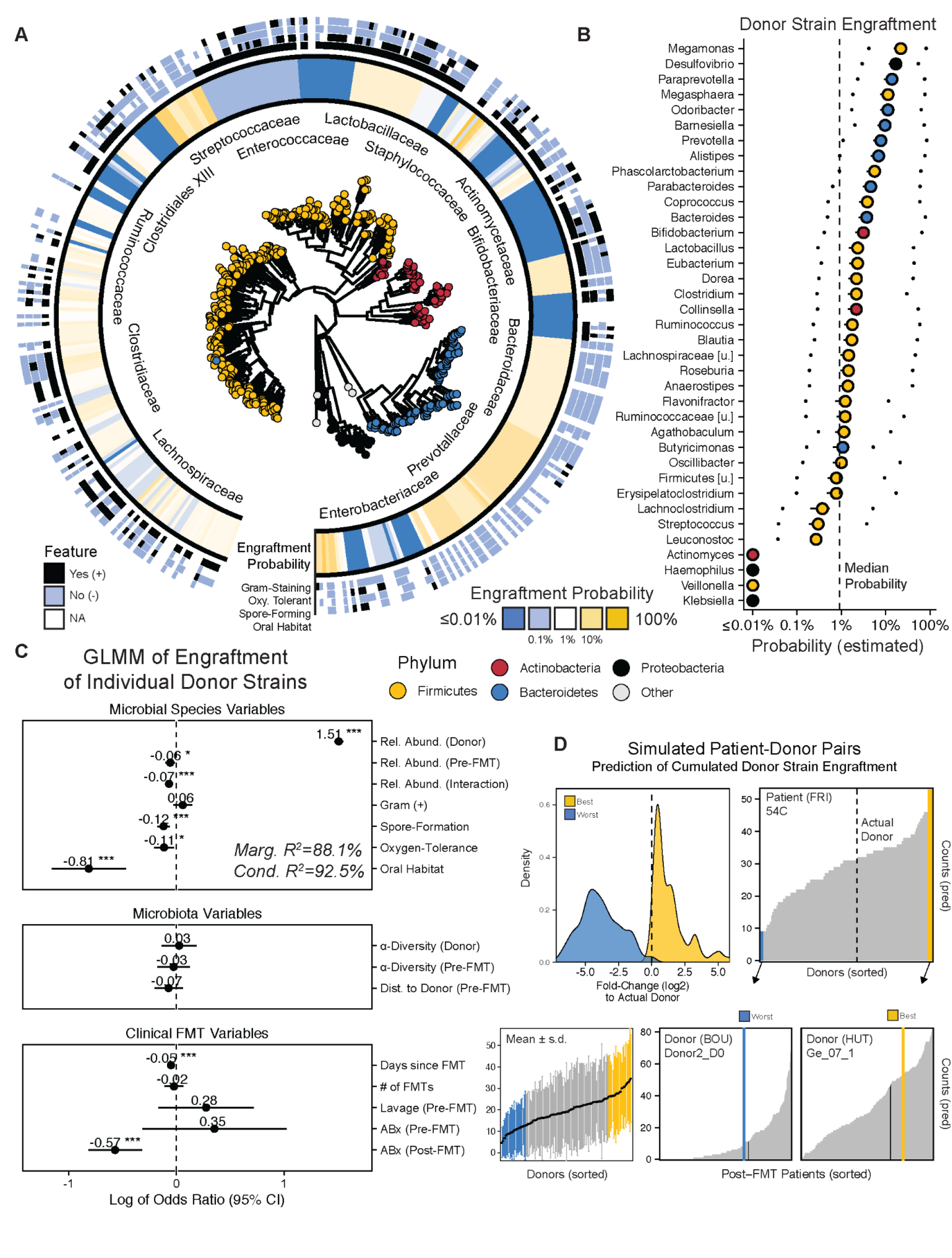
Donor strain engraftment probabilities for individual strains. **A)** Estimated donor strain engraftment probabilities in relation to phylogeny and microbial species features (Gram-staining, spore formation, oxygen-tolerance, oral habitat), using the generalized linear mixed-effects model (GLMM). **B**) Median donor strain engraftment probabilities for different genera, together with the estimated minimum and maximum probabilities, when using the lowest and highest species relative abundances that were detected in the meta-cohort as alternative input variables to the model. **C**) Species features, (patient and donor) microbiota parameters, and clinical FMT variables with relevance for the engraftment of individual donor strains based on the GLMM. Asterisks denote significance thresholds: *p≤.01, **p≤.001, ***p≤.0001. **D**) Total numbers of donor-derived strains in post-FMT patients, based on GLMM predictions for individual donor strains, vary substantially depending on recipient/donor pairing. *Top left*: They deviate ± 5-fold (log2) for the worst (blue) and best (yellow) simulated donor pair, relative to the actual recipient/donor pairs. *Bottom left*: Variations between different recipient/donor pairs for one donor (y-axis range) generally exceed variations between different donors (x-axis range). Black dots and bars: mean values ± SD. *Top right, bottom middle, and right*: Using patient 54C as an example, the predicted engraftment varies between <10 and >40 strains for different donors. But the worst (blue) and best (yellow) donors for this patient are predicted to also result in a broad range of engrafted donor strains in pairings with other patients. Dark lines indicate observations from actual recipient/donor pairings.

### Engraftment prediction for individual donor strains based on recipient and donor information

There is substantial interest and a strong rationale to apply personalized approaches to improve the clinical performance of FMT and induce health benefits not only in rCDI and IBD but also in other gastrointestinal disorders (Benech and Sokol 2020). To explore the potential of FMT personalization, we assessed the effects of different - simulated - recipient/donor pairings on donor strain engraftment. For this, we predicted the engraftment of all detected donor strains for all possible patient/donor combinations from the FMT meta-cohort. First, donor strain engraftment probabilities were re-estimated for each FMT case based on the real input variable set, except that the species relative abundance in the donor was replaced with the highest or lowest species relative abundance that was detected in any of the other donors from the meta-cohort (Fig. 5B). Under this model, donor strain engraftment for many genera became substantially more or less likely, in particular for genera from the phylum *Firmicutes* (Fig. 5B). For example, while the genus *Ruminococcus* carried a median donor strain engraftment probability of 1.75% across the entire FMT meta-cohort, it was estimated that this probability could be reduced to 0.23% or increased to 58.9% with the highest and lowest detected cumulative relative abundance of *Ruminococcus* species in any of the other FMT donors, respectively. These simulations are noteworthy, as they could inform FMT-based therapeutic developments on the basis of *in vitro*-supplemented donor samples with select microbial cultures.

Next, we predicted FMT outcomes on the basis of the total number of engrafted donor strains that would be expected for all possible patient/donor combinations from the meta-cohort. Here, we compared patient/donor pairs based on the total number of donor strain engraftment events that were predicted based on all detected donor species and actual recipient and donor microbiota compositions. For the subset of real combinations among the simulated patient/donor pairs, we observed a strong correlation between predicted and detected donor strain engraftment numbers (r=0.94, p<0.0001), demonstrating a good fit of our mixed model to the data. Across the entire meta-cohort, the predicted total number of engrafted donor strains per patient varied in a range of log2(±5)-fold between the worst and best-engrafting donors relative to the actual donor (Fig. 5D). Donors that performed poorly in one recipient (<10 donor-derived post-FMT strains) were predicted to produce substantially larger numbers of engrafted strains in other patients (>50 donor-derived post-FMT strains) or *vice versa* (examples shown in Fig. 5D). In general, recipient/donor pairings accounted for more variation in the predicted number of engrafted donor strains than the use of different donors (Fig. 5D). Our findings show that donor strain engraftment can be predicted based on recipient and donor microbiota information. These findings provide the basis for personalized applications of FMT with the goal to maximize donor microbiota engraftment in patients by pairing them with specific donors.

### Strain engraftment is linked to clinical response in ICI

Cure rates of ∼90% after FMT have been reported for rCDI in patients after repeated antibiotic treatment failure (Khoruts, Staley, and Sadowsky 2021), which, based on our models, would induce dysbiosis in patients before and increase donor strain engraftment after FMT. Consequently, rCDI patients from our meta-cohort, all of which resolved symptoms after FMT, consistently acquired large donor-derived strain proportions after the treatment (Fig. 3C), suggesting that FMT outcome in this patient population may be dependent on donor microbiota engraftment. However, this could not be systematically analyzed, as fecal metagenomes from failed treatment cases are scarce or lack the sequencing depth that is required for strain-level microbiota profiling (Kazemian et al. 2020). Therefore, we focused the analysis on a subset of two IBD and two ICI studies from the meta-cohort for which clinical response information was available (Fig. 6). In general, variations in donor strain engraftment between studies and study subgroups, which, based on our mixed model, were due to different patient pretreatment and FMT protocols (ABx, no. of FMTs), by far exceeded differences between responders and non-responders from the same study. For ICI, responders from one study (Baruch et al. 2021) carried larger proportions of donor-derived strains and these strains represented larger relative abundance fractions than non-responders. In responders from the second ICI study (Davar et al. 2021) the same trend but no significant difference was observed, suggesting that additional, larger studies will be needed to determine the role of donor microbiota engraftment for the induction of a clinical response to ICI in melanoma patients. In contrast, neither consistent differences within nor across the two analyzed IBD studies or their subgroups were observed between responders and non-responders with respect to donor microbiota engraftment (Fig. 6) or α/β-diversity microbiota parameters (Suppl. Fig. S4). In summary, the impact of donor microbiota engraftment on the clinical response to FMT remains unclear and may be disease-dependent. However, the available data indicate that untargeted FMT, i.e. with the goal to maximize absolute donor microbiota engraftment in the patient, may be more relevant for rCDI than IBD and ICI-refractory melanoma. In the latter patient populations, personalized FMT approaches to induce engraftment of specific donor strains as illustrated in our simulations may be more important.

**Fig. 6.**
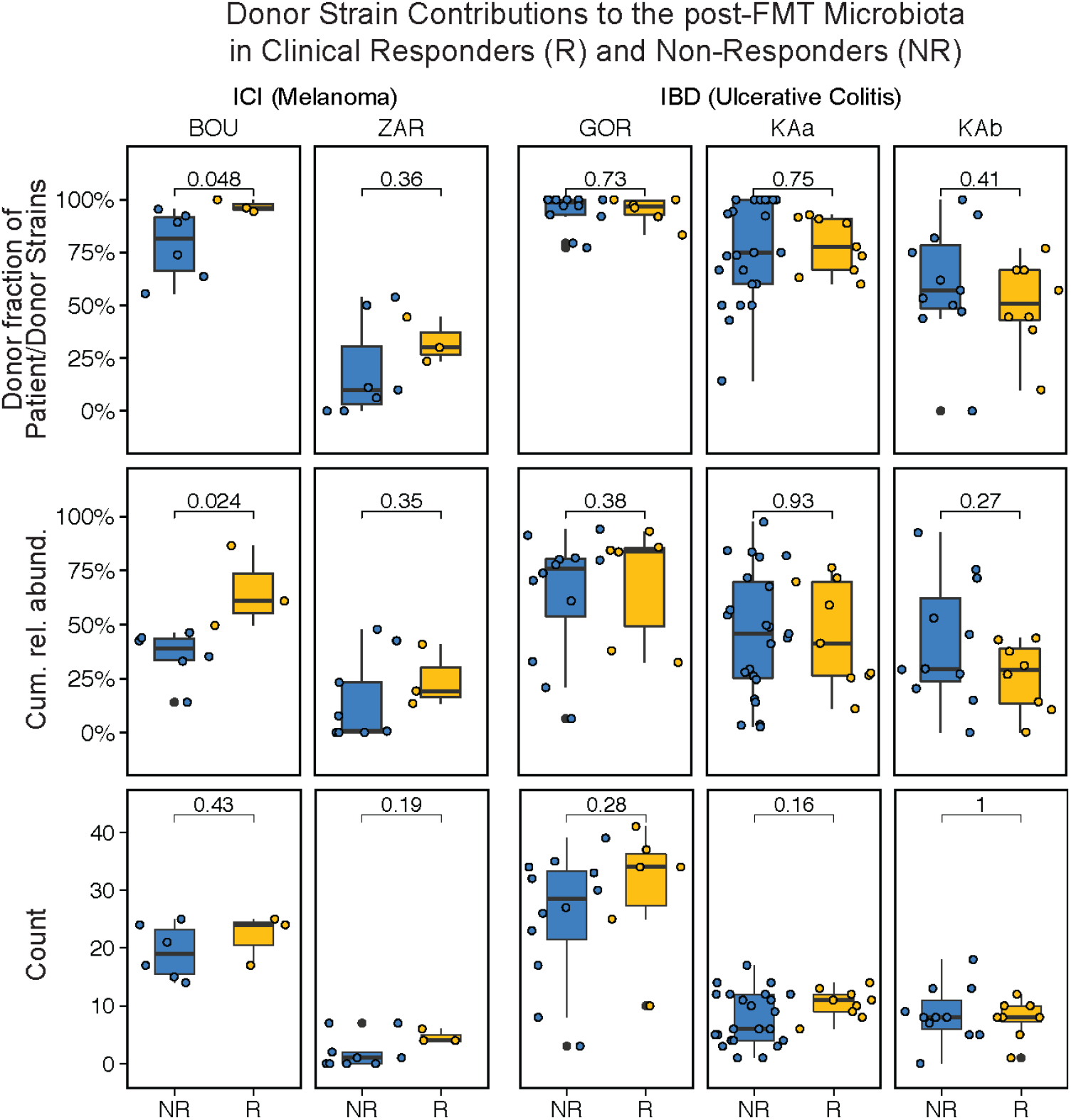
Donor microbiota engraftment and clinical response to FMT. Responders (R, yellow) and non-responders (NR, blue) from two FMT trials to overcome resistance to anti-PD-1 therapy in ICI-refractory melanoma patients and two FMT trials to induce remission in IBD patients were compared based on donor-derived strain fractions (of recipient and donor-derived strains), cumulative relative abundances of species represented by donor-derived strains and total numbers of donor-derived strains in post-FMT samples (last available sample within ≤4 months after FMT). See Methods for a description of R/NR criteria. IBD patients from the two study branches of Paramsothy et al., which applied different FMT protocols resulting in different levels of donor microbiota engraftment (Fig. 3C), were compared separately.

## DISCUSSION

FMT is both a promising therapeutic avenue for microbiome-associated pathologies as well as an experimental tool to establish a causal role of the gut microbiota in human disease. For rCDI, FMT achieves a ∼90% cure rate (Khoruts, Staley, and Sadowsky 2021) and high levels of post-FMT donor microbiota engraftment in patients (>50% donor-derived strains, Fig. 3C), not only attesting to the efficacy of a now commonly used therapeutic option but also allowing for the causal inference of the gut microbiota in a human pathology (Walter et al. 2020). More modest and inconsistent clinical benefits have been reported for the treatment of IBD (Danne, Rolhion, and Sokol 2021), ICI-refractory melanoma (Woelk and Snyder 2021), and MetS (Hanssen, de Vos, and Nieuwdorp 2021) with FMT. However, donor microbiota engraftment in these patient populations had previously also not been comprehensively characterized, compared and correlated to individual patient parameters, FMT modalities or clinical outcome parameters. The therapeutic potential of FMT for these and other microbiota-associated disorders therefore remains mostly unclear. Moreover, the ecological principles that govern the post-FMT patient microbiota assembly process, as well as their relationship with underlying medical conditions and clinical FMT modalities are insufficiently understood. We present the first detailed characterization of donor strain engraftment in a diverse FMT-treated patient cohort with a heterogeneous background of medical conditions, microbiota compositions and FMT procedures, based on a meta-analysis of newly generated and available metagenomes with our recently introduced SameStr tool for the detection of donor-derived strains in post-FMT patient samples (Podlesny and Fricke 2020)(Podlesny et al., submitted). Using generalized linear mixed-effects models, we measured associations between donor strain engraftment and the recipient and donor microbiota (ɑ/β-diversity, dysbiosis, microbial taxonomy and function), patient background (medical condition, preparation for FMT), and clinical FMT modalities (no. of FMTs). As a result, we provide the first general models for post-FMT microbiota assembly, which allow for the prediction of patient and strain-specific donor microbiota engraftment based on case-specific microbiome parameters. We determine clinically modifiable factors as targets for FMT outcome optimization, and we identify distinct clinical goals for FMT optimization, i.e. either (i) to maximize broad, untargeted engraftment of the donor microbiota after FMT with antibiotic patient pretreatment, with likely clinical benefits for the treatment of rCDI, or (ii) to increase the engraftment probability of specific donor strains based on recipient/donor matching. The latter scenario supports development of personalized applications of FMT in different pathologies within a framework of precision medicine.

A key finding of our study is that donor strain engraftment after FMT is strongly dependent on recipient microbiota composition and dysbiosis. In rCDI, where dysbiosis is rampant (Fig. 1) and linked to taxonomic (e.g. lower ɑ-diversity, altered β-diversity) and functional (e.g. increased relative abundance of oral and oxygen-tolerant species) microbiota disruption (Fig. 2), FMT not only resolves dysbiosis but also results in contributions of 60-90% donor strains to the post-FMT patient microbiota (Fig. 3). In agreement with previous studies that did not detect strong dysbiotic patterns in IBD and obesity (Lloyd-Price et al. 2019; Duvallet et al. 2017; Sze and Schloss 2016), especially when controlling for confounding host variables (Vujkovic-Cvijin et al. 2020; Stanislawski et al. 2019), we demonstrate the absence of detectable dysbiosis in other FMT-treated medical conditions, including IBD, MetS, and ICI. However, dysbiosis was inducible by patient pretreatment with antibiotic cocktails containing vancomycin (specific for Gram-positive bacteria) and to a lesser extent with antibiotics specific for Gram-negative bacteria. Similar to antibiotic treatment (Modi, Collins, and Relman 2014), bowel lavage results in reduced intestinal microbiota loads and altered fecal microbiota compositions (Jalanka et al. 2015) with consequences for colonization resistance to invading pathogens (Litvak and Bäumler 2019). Accordingly, donor-derived strain contributions to the post-FMT microbiota were modest (<50%), unless patients underwent microbiota depletion by antibiotic treatment and bowel lavage or received extensive repeated FMTs (>40 enemas) (Fig. 3). Our findings therefore point to the disruption of the patient microbiota colonization resistance as a clinical target for FMT optimization. In light of the collateral effects of antimicrobial drugs on resistance development, microbiota ecology (Modi, Collins, and Relman 2014), and host health (Filippone, Kraft, and Farber 2017) alternative strategies for patient pretreatment should be further explored, e.g. through dietary interventions, such as fiber depletion (Sonnenburg et al. 2016) or caloric restriction (von Schwartzenberg et al. 2021). Importantly, our findings suggest that with microbiota depletion in preparation for FMT extensive donor microbiota engraftment can be achieved independently of the underlying medical conditions of the patients, potentially broadening the spectrum of microbiota-associated diseases that could be treatable by FMT.

Our findings provide important insights into the ecological principles of post-FMT microbiota assembly. First, donor and recipient strains from the same species rarely coexisted in post-FMT patients from our meta-cohort, independently of disease context, dysbiosis, or FMT procedure, indicating a competitive exclusion between closely related strains with resource niche overlaps in agreement with coexistence theory (Letten, Hall, and Levine 2021). Second, post-FMT donor microbiota engraftment was reduced in non-dysbiotic patients (i.e. without ABx pretreatment), indicative of an increased competitive ability of resident recipient strains in undisturbed microbiomes, in line with priority effects that benefit earlier microbiome colonizers over later immigrating strains (Fukami 2015). Third, antibiotic patient pretreatment shifted FMT outcomes towards increased donor microbiota engraftment, effectively overcoming priority effects, as recently demonstrated for consecutive strain colonization experiments in gnotobiotic mice (Munoz et al. 2020). These ecological interactions provide a mechanistic framework that explains the generalizability of our predictive models for patient and strain-specific donor microbiota engraftment across an extremely heterogeneous clinical meta-cohort.

We and others identified a strong microbiota signal of dysbiosis in rCDI patients, which is resolved after FMT (Song et al. 2013; Weingarden et al. 2015), leading to substantial donor microbiota engraftment (Smillie et al. 2018)(Podlesny et al., submitted), providing strong evidence for a causal role of the microbiota and the clinical potential of FMT-induced donor microbiota engraftment as a treatment. A causal role of dysbiosis for pathologies other than rCDI has not been clearly established, and IBD, ICI, and MetS were not associated with a detectable dysbiosis in our meta-cohort. Consequently, the clinical value of FMT for the treatment of these conditions remains unclear and previous FMT studies lacked the taxonomic resolution to link donor strain engraftment to clinical outcomes. We compared strain engraftment in responders and non-responders for a total of four IBD and ICI studies for which clinical outcome data was available. We did not observe differences in donor strain engraftment in IBD responders and non-responders, and although donor microbiota engraftment rates varied substantially between studies, they produced similar clinical outcomes (Fig. 6). However, we observed a significantly larger contribution of donor-derived strains to the post-FMT microbiota of ICI-refractory melanoma patients that responded to FMT with an objective reduction in tumor size after anti-PD-1 treatment in one trial and saw a similar trend in a second trial, despite low patient numbers in both studies (n≤15). These findings suggest a role of donor microbiota engraftment for the FMT-induced response to ICI therapy, which should be explored. They further highlight the utility of our strain-level microbiota profiling approach to quantify the impact of FMT on post-FMT microbiota assembly. Future studies should expand our analysis and use strain-level microbiota analysis to quantify both broad untargeted donor microbiota engraftment and to identify specific donor strain engraftment or recipient strain replacement events in relation to clinical outcome parameters of FMT. Such strains could be used in follow-up experiments, e.g. in animal models of the respective pathology, as a first step to prove a causal microbiota involvement in disease etiology (Walter et al. 2020).

Studies are beginning to outline personalized FMTs strategies within the conceptual framework of precision medicine by drawing attention to both recipient and donor microbiome features to predict and optimize clinical FMT outcomes (Benech and Sokol 2020). The post-FMT microbiota assembly models that we have developed based on donor and recipient variables showed that the number of engrafted donor strains can be substantially increased (estimated: >10-fold) by pairing recipients with specific donors. Our models can further be applied to predict the engraftment of specific donor strains (e.g. from the genus *Blautia*), providing the theoretical basis for precision microbiota modulation therapies by targeting donor strains with desirable genetic traits (e.g. lantibiotic-producing, resistance to vancomycin-resistant Enterococcus-restoring, *Blautia producta* strains (S. G. Kim et al. 2019)) for introduction into a patient’s microbiota. Although not experimentally validated yet, our findings offer the intriguing perspective to improve FMT outcomes for previously tested clinical applications and open up new opportunities for FMT-based precision microbiome modulation therapies. In practice, personalized FMT strategies that should be further tested could involve fecal sample selection from extensive door stool banks, as well as supplementation of fecal samples with specific strains, as strain abundance in the donor sample, or dosage effects, were most predictive of strain engraftment in our model.

Our study has several limitations, resulting from the extremely heterogeneous FMT meta-cohort involving different medical conditions, patient preparations, FMT procedures, single/multi-donor combinations, clinical pre- and post-FMT patient metadata, and sampling timelines. Most trials involved small patient numbers, which were sampled at different and inconsistent time points, often involving only a single sample collected from patients before pretreatment. Therefore, although or models identified strong and distinct effects of bowel lavage and antibiotic patient pretreatment on donor microbiota engraftment, these and other comparisons (e.g. on the impact of multiple FMTs) were only based on patient subsets from out meta-cohort, and the temporal trajectories of the post-FMT assembly process were not characterized in detail. Therefore, the use of an extensive heterogeneous meta-cohort was likely instrumental in the development of robust predictive models and the identification of what appear to be the universal drivers of post-FMT microbiota assembly. Our retrospective meta-analysis does not allow for the validation of our predictions for donor strain engraftment, which would be needed to test our capacity for precise recipient/donor matching-based microbiota modulation. Similarly, the clinical assessment of FMT outcomes, which we have broadly studied in the context of broad, untargeted donor microbiota engraftment, was not the focus of the study and should be further analyzed.

In summary, our findings broadly characterize the contributions of donor-derived strains to the post-FMT patient microbiota across a diverse set of patient, microbiome and clinical conditions. They suggest a major impact of adjustable patient pretreatment modalities on donor strain engraftment and present generalizable predictive models for patient and strain-specific donor microbiota engraftment that are in agreement with ecological theory. They further illustrate the theoretical potential for personalized FMT applications through fecal supplementation with select strains and recipient/donor matching to increase the engraftment probability of specific strains. With this work, we lay the groundwork for future developments of precision microbiota modulation therapies.

## MATERIALS AND METHODS

### Study design

The objective of this study was to characterize the microbiota of FMT-treated patients with different medical conditions (“meta-cohort”) and their donors and to apply strain-level microbiota profiling in order to determine donor-derived strain contributions to post-FMT microbiota assembly. This information was used to inform predictive models to describe the post-FMT microbiota assembly process and to determine the role of recipient and donor microbiota features, taxonomic and ecological microbiota parameters, and clinical modalities of patient preparation and FMT for donor strain engraftment.

### Study cohort

We assembled a comprehensive meta-cohort of 245 distinct clinical cases in which FMT was used to modulate the microbiota of patients with different medical conditions (Fig. 1A). Published across thirteen distinct studies, FMT was used to treat recurrent *Clostridioides difficile* infection (rCDI) (Watson et al. 2021; Smillie et al. 2018; Song et al. 2013)(Podlesny et al., submitted), type 2 diabetes mellitus, obesity and metabolic syndrome (MetS) (Li et al. 2016; Ng et al. 2021; Wilson et al. 2021), the inflammatory bowel diseases Crohn’s disease and ulcerative colitis (IBD) (Kong et al. 2020; Paramsothy et al. 2019; Kump et al. 2018), to eradicate multidrug-resistant *Enterobacteriaceae* carriage (MDR) (Bar-Yoseph et al. 2020; Leo et al. 2020), and to induce response to anti–PD-1 immunotherapy in immune checkpoint inhibitor therapy-refractory melanoma patients (ICI) (Davar et al. 2021; Baruch et al. 2021). Fecal metagenomic shotgun sequence data were included from a total of 1232 samples obtained from patients before and (often multiple times) after treatment (Fig. 1E), as well as from stool donors (Table S1, S2). Metadata were obtained from the supplementary information provided with each publication or from the authors.

Clinical response information was obtained from the original publications and defined as follows: For ICI patients (see Fig. 1B in both Baruch et al. and Davar et al.), responders (R) experienced an objective response to treatment as per Response Evaluation Criteria in Solid Tumors (BOU: iRECIST (Seymour et al. 2017), ZAR: RECIST v1.1 (Eisenhauer et al. 2009)), indicated by a tumor regression of at least 30% compared to baseline, whereas non-responders (NR) showed progressive disease with an increase in tumor size of at least 20%. For IBD patients, responders (R) went into remission (Mayo score (Lewis et al. 2008): ≤2), all other IBD patients were classified as non-responders (NR).

### Reference cohort

To compare microbiota composition metrics against a healthy control cohort, fecal metagenomic shotgun sequence data from subjects that had not reported conditions that would suggest extensive medication or strong microbiota perturbations were obtained through the curatedMetagenomicsData package (Pasolli et al. 2017). For each subject, sequence data downloaded from NCBI’s Sequence Read Archive were concatenated in case of multiple available accessions. We additionally collected preprocessed MetaPhlAn3 species-level taxonomic relative abundance profiles that were made available with bioBakery 3 (Beghini et al. 2021). In sum, 739 samples from nine publicly available datasets (Table S4) were used for the reference cohort (Human Microbiome Project Consortium 2012; Raymond et al. 2016; Zeller et al. 2014; Yu et al. 2017; Feng et al. 2015; Vogtmann et al. 2016; Thomas et al. 2019; Wirbel et al. 2019; Yachida et al. 2019).

### Quality control and preprocessing of metagenomic shotgun sequence data

All raw paired-end metagenomic sequence reads were processed with KneadData v0.6.1 to trim sequence regions with base call quality below Q20 within a 4-nucleotide sliding window and to remove reads that were truncated by more than 30% (SLIDINGWINDOW:4:20, MINLEN:70). To remove host contamination, trimmed reads were mapped against the human genome (GRCh37/hg19) with bowtie2 v2.2.3 (Langmead and Salzberg 2012). Output files consisting of surviving paired and orphan reads were concatenated and used for further processing.

### Taxonomic and functional community composition analysis

Preprocessed sequence reads from each sample were mapped against the MetaPhlAn clade-specific marker gene database (mpa_v30, 201901, Table S5) using MetaPhlAn3 v3.0.7 (Beghini et al. 2021). Relative abundances of species-level taxonomic profiles were centered-log ratio (clr) transformed and used for principal component analysis with FactoMineR v2.4 (Lê, Josse, and Husson 2008). Density ridgeline plots were generated with the ggridges package v0.5.3 in R v3.6.1. Shannon Index and Bray-Curtis dissimilarity were determined with vegan v2.5.7 (diversity function), and the UniFrac distance with phyloseq v1.28.0 (UniFrac function) using the phylogenetic tree published along MetaPhlAn3. Functional metadata on bacterial species (Table S6) were aggregated from different publications (Browne et al. 2016; Vatanen et al. 2019), the List of Prokaryotes according to their Aerotolerant or Obligate Anaerobic Metabolism (OXYTOL 1.3, Mediterranean institute of infection in Marseille), bacDive (Reimer et al. 2019), FusionDB (Zhu et al. 2018), The Microbe Directory v2.0 (Sierra et al. 2019), and the expanded Human Oral Microbiome Database (Escapa et al. 2018). For each sample, the cumulative relative abundance of taxa that were associated with oxygen tolerance or an oral habitat was determined (Fig. 2). The Microbial Dysbiosis Score was calculated as the log-ratio of the cumulative relative abundance of taxa which were previously positively and negatively associated with pediatric Crohn’s Disease (Gevers et al. 2014).

To detect significant differences in patients and donors relative to healthy controls (Fig. 2) with respect to microbiota composition (ɑ/β-diversity), dysbiosis and bacterial species lifestyles (oral habitat, oxygen tolerance) generalized linear mixed-effects models (GLMMs) were used, which were calculated with the glmer function (binomial link) in lme4 v1.1.27 (Bates et al. 2015) or the lmer function from lmerTest v3.1.3 (Kuznetsova, Brockhoff, and Christensen 2017). For each study and metric, the sample type (pre-FMT^ABx-^, pre-FMT^ABx+^, post-FMT, donor, control) was incorporated as a fixed effect, using samples from the control cohort as a reference. Only post-FMT patient sample data were included that were collected at least five days after FMT. We controlled for study effects in the control cohort and repeated post-FMT patient sampling by including study and case as random effects. The resulting model outputs are tabulated in Table S7.

### Detection of shared strains with SameStr

To track bacterial strains in distinct biological samples, we used the SameStr tool from our group (Podlesny and Fricke 2020)(Podlesny et al., submitted), which leverages the clade-specific MetaPhlAn markers to resolve within-species phylogenetic sequence variations. Briefly, MetaPhlAn3 marker alignments were converted to single nucleotide variant (SNV) profiles, extensively filtered, merged, and compared between metagenomic samples based on the maximum variant profile similarity (MVS) to detect strains that were shared between samples. In contrast to StrainPhlAn, which uses the major allele at every position in the alignment (consensus sequence), SameStr’s MVS-based approach evaluates the co-occurrence of all four possible nucleotide alleles between overlapping alignment sites of two samples, including polymorphic sites (≥10% allele frequency), which can result from multiple strains representing the same species. This allows for the detection of sub-dominant shared strains or coexisting recipient and donor strains from the same species. Shared strains were called if species alignments between metagenomic samples overlapped by ≥5 kb and with an MVS of ≥99.9%. The SameStr program and further documentation is available at GitHub: https://www.github.com/danielpodlesny/SameStr.git.

### Identification of recipient and donor-derived strains in post-FMT patients

For each post-FMT patient sample, recipient and donor-derived taxa were determined based on shared species and strains with pre-FMT recipient and donor samples. Recipient or donor-derived or coexisting strains were identified as shared exclusively between post-FMT and pre-FMT patient, or between post-FMT patient and donor, or between post-FMT and pre-FMT patient and donor samples, respectively. Analogously, recipient or donor-derived species were exclusively shared between post-FMT and pre-FMT patient, or between post-FMT patient and donor samples, respectively. In several cases, multiple available samples from the same donor or individual samples from multiple donors that were used for pooled FMTs were combined. For Wilson et al. (OSU), individual donor samples that the authors had combined in batches for FMT were combined. For Li et al. (BOR), which included data from a single donor sampled at three distinct time points without providing information about their specific use for distinct patients, donor samples were combined, as described in the original publication. For Kump et al. (GOR), which included repeated FMT, all available sequence data for each donor were combined across treatment rounds. Ng et al. (CHA) did not disclose sample metadata in the publication, including information about patient/donor pairing and patient assignments to different treatment groups (FMT alone, FMT with lifestyle intervention, or sham treatment), and this information was not made available upon requests to the authors. For this study, concatenated sequence files of all five donor samples were used and sham cases identified and excluded based on the lack of shared strains with any post-FMT sample (see Suppl. Fig. S2). In case of pooled donor sequence data, mean values of all relevant microbiota metrics (e.g. ɑ/β-diversity) were used in our models. Baruch et al. (BOU) published fecal metagenomes from patients before but not after antibiotic pretreatment. To include this dataset in our models, we imputed pre-FMT microbiota metrics with the mean values that were observed in all antibiotically pretreated patients. We also tested our model with the BOU data, which had only a minor effect on the model predictions (Suppl. Fig. S3)

### Post-FMT assembly models

Generalized linear mixed-effects models (GLMMs) were used to estimate the effects of recipient and donor microbiota parameters and clinical modalities on the post-FMT microbiota assembly process. Separate GLMMs were used to determine the role of these parameters for donor-derived strain fractions in individual patients after FMT (Fig. 4) and of these and additional, species-specific parameters for the engraftment of individual donor strains (Fig. 5).

Overall donor microbiota engraftment (Fig. 4) was calculated as the donor-derived strain fraction of the total number of donor-derived and recipient-derived strains per post-FMT sample. By calculating strain fractions, our donor microbiota engraftment metric reduces confounding effects, as variable sequencing depths affect the total number of detectable strains per sample. As a consequence, the ratio of donor-derived to recipient and donor-derived strain numbers is reflective of the degree to which the recipient microbiota is replaced by the donor microbiota after FMT, whereas donor-derived strain numbers alone would not be able to differentiate microbiota replacement from microbiota expansion, i.e. an engraftment of donor strains on top of persisting recipient strains. Donor-derived post-FMT strain fractions were modeled across the entire meta-cohort by incorporating centered and scaled microbiota and clinical parameters as fixed effects and by controlling for repeated patient sampling and study effects by including case and study as random effects. Marginal effects were calculated with ggeffects v1.1.0 (ggpredict function) and model coefficients visualised in a forest plot with sjPlot v2.8.8. The resulting model outputs are tabulated in Table S8-S9.

We expanded our model to estimate the relevance of relative abundances and functional features (Gram stain, spore formation, oxygen tolerance, oral habitat) on the engraftment probabilities of individual strains in our meta-cohort (except Ng et al. for which patient/donor pairing information was not available). Using all species that were detected in a donor and provided only with donor and pre-FMT patient microbiota information, the model predicted whether the corresponding donor-derived strain would be detected in the post-FMT patient or not. Functional features were coded on the species level as +1 (yes, positive) and −1 (no, negative), replacing missing values with each feature’s mean frequency across all species (in order to avoid an influence on the model). Since some donors were used to treat multiple patients, in addition to repeated sampling and study effects, we also controlled for donor-specific influences with random effects in our model. The adjusted engraftment probability of each genus (Table S9) is shown in the context of a phylogenetic tree (Fig. 5A, ggtree v1.16.6) that is annotated with lifestyle features for each species, and as a point range (± s.e.), with probabilities of each genus additionally conditioned on the minimum and maximum relative abundance observed within the donor population (Fig. 5B).

The GLMM for the engraftment of individual strains closely fit the underlying data, both for predicted donor strain engraftment events of individual genera and aggregating total numbers of engrafted strains (r=0.94, p<.0001). Therefore we used the model to simulate FMT outcomes for other recipient/donor pairs from the meta-cohort. For these simulations, species-level taxonomic compositional profiles of recipient and donor samples, as determined with MetaPhlAn3, were used as input for the GLMM and the predicted total number of engrafted donor strains (based on individual predictions for each donor strain) were aggregated for each patient and time point and compared to actually detected numbers from the real recipient/donor pairs by calculating (log2) fold-changes (Fig. 5D

## Data Availability

Shotgun metagenomic data from the UC Study Cohort by Kump et al. will be deposited and made available at the European Nucleotide Archive.

## Acknowledgments

**Funding:** D.P. and W.F.F. received funding from the German Research Foundation (DFG, Deutsche Forschungsgemeinschaft) under SPP 1656 (Project no. 316130265).

## Author Contributions

Conceptualization and Methodology, D.P., J.W. and W.F.F.; Writing of the Original Draft, D.P., J.W. and W.F.F.; Review & Editing, D.P., J.W. and W.F.F.; Software, Validation, and Formal Analysis, D.P.; Resources, M.D., S.P., N.O.K., and G.G.; Data Curation, D.P., M.D., S.P.; Funding Acquisition, W.F.F.

## Supplementary Materials

Table S1. Sample and case metadata.

Table S2. WGS accession identifiers.

Table S3. WGS QC data.

Table S4. Control Cohort accessions.

Table S5. MetaPhlAn3 species-level taxonomic profiles.

Table S6. Species-level metadata on microbe lifestyle.

Table S7. Donor and patient contributions to the post-FMT microbiota.

Table S8. Results for GLMM of Donor Strain Engraftment in Individual Patients

Table S9. Results for GLMM of Engraftment of Individual Donor Strains

## Supplemental Figures

**Fig. S1.**
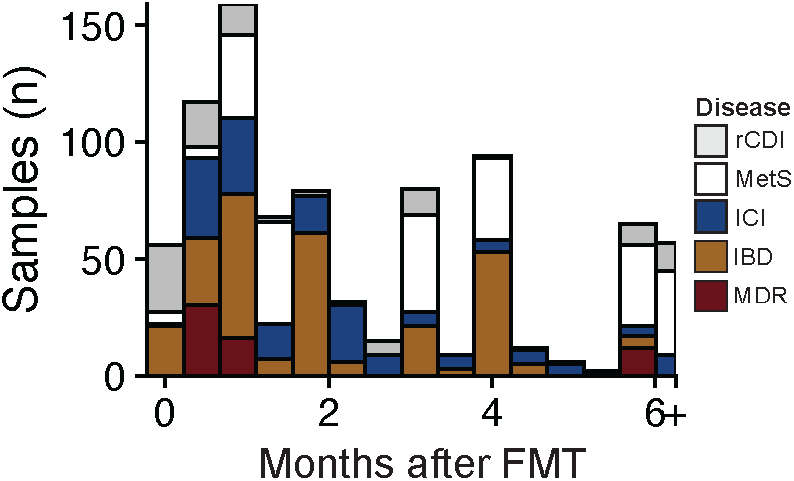
Available sample distribution across different post-FMT time points. The histogram is colored by disease categories.

**Fig. S2.**
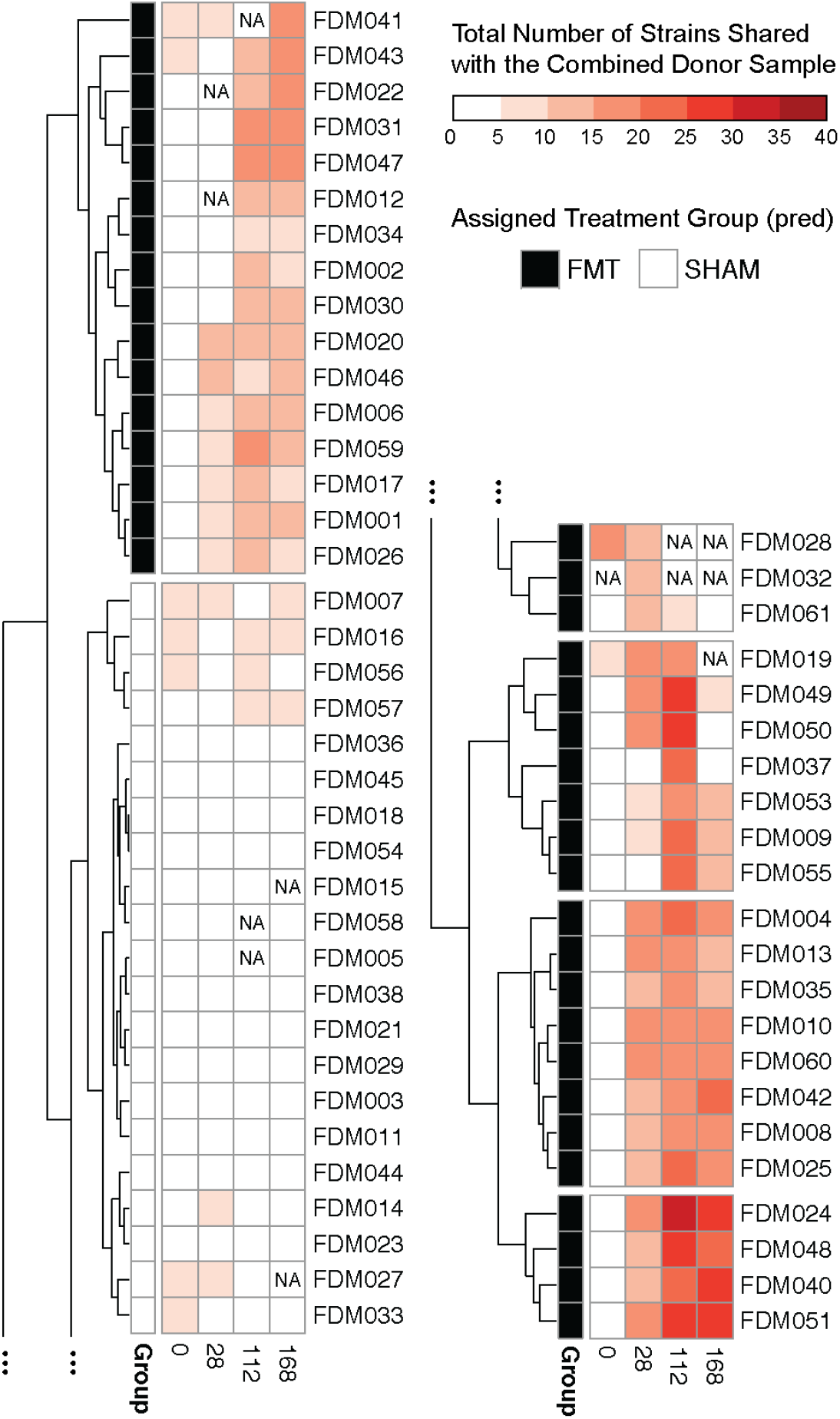
Clustering of all samples from Ng et al. based on shared strain profiles in order to determine the assignment of patients to the FMT or sham treatment groups. Hierarchical clustering of strain-sharing profiles between a concatenated donor sample (combined from five published donor metagenomes) and each patient from Ng et al. at one pre-FMT (0) and three post-FMT (28, 112, 168) time points highlights 21 patients whose samples share very few (mostly <5) strains with any donor sample. Since treatment assignments were not disclosed by Ng et al., these 21 patients were excluded from the analysis as they most likely belong to the sham treatment group of the study.

**Fig. S3.**
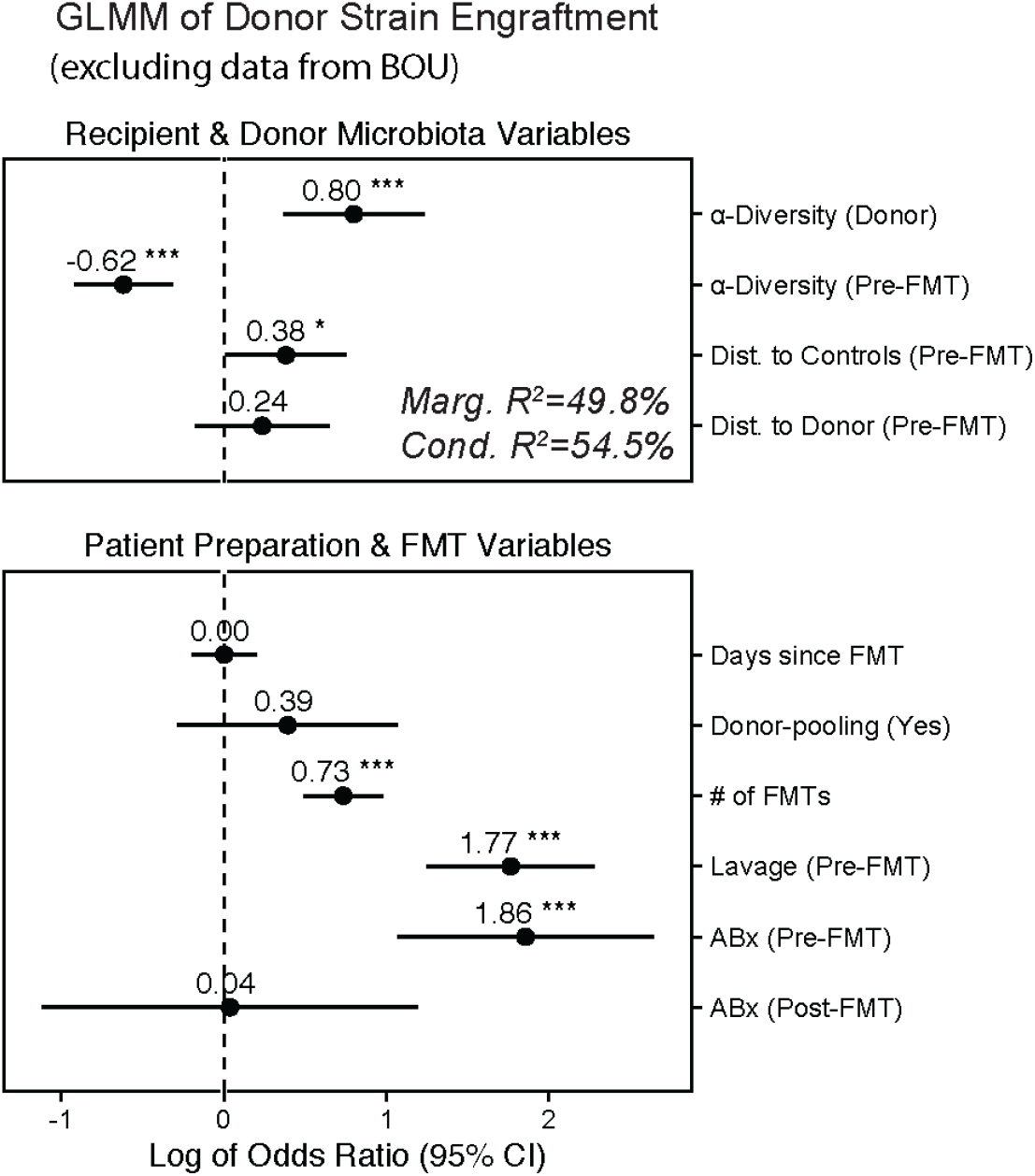
Related to Fig. 4. GLMM of donor strain engraftment excluding data from Baruch et al. Baruch et al. (BOU) collected pre-FMT samples before pretreatment with antibiotics. As for this dataset patient ɑ-diversity and β-diversity to controls and donors was unavailable for the pre-FMT^ABx-^ sample, which would be most relevant for our model, the mean of those values that were observed in all other antibiotically treated pre-FMT patients was used instead to generate the model shown in Fig. 4. Here we generated a similar model without the data from Baruch et al., showing very similar results (odds ratio and significance) as in Fig. 4.

**Fig. S4.**
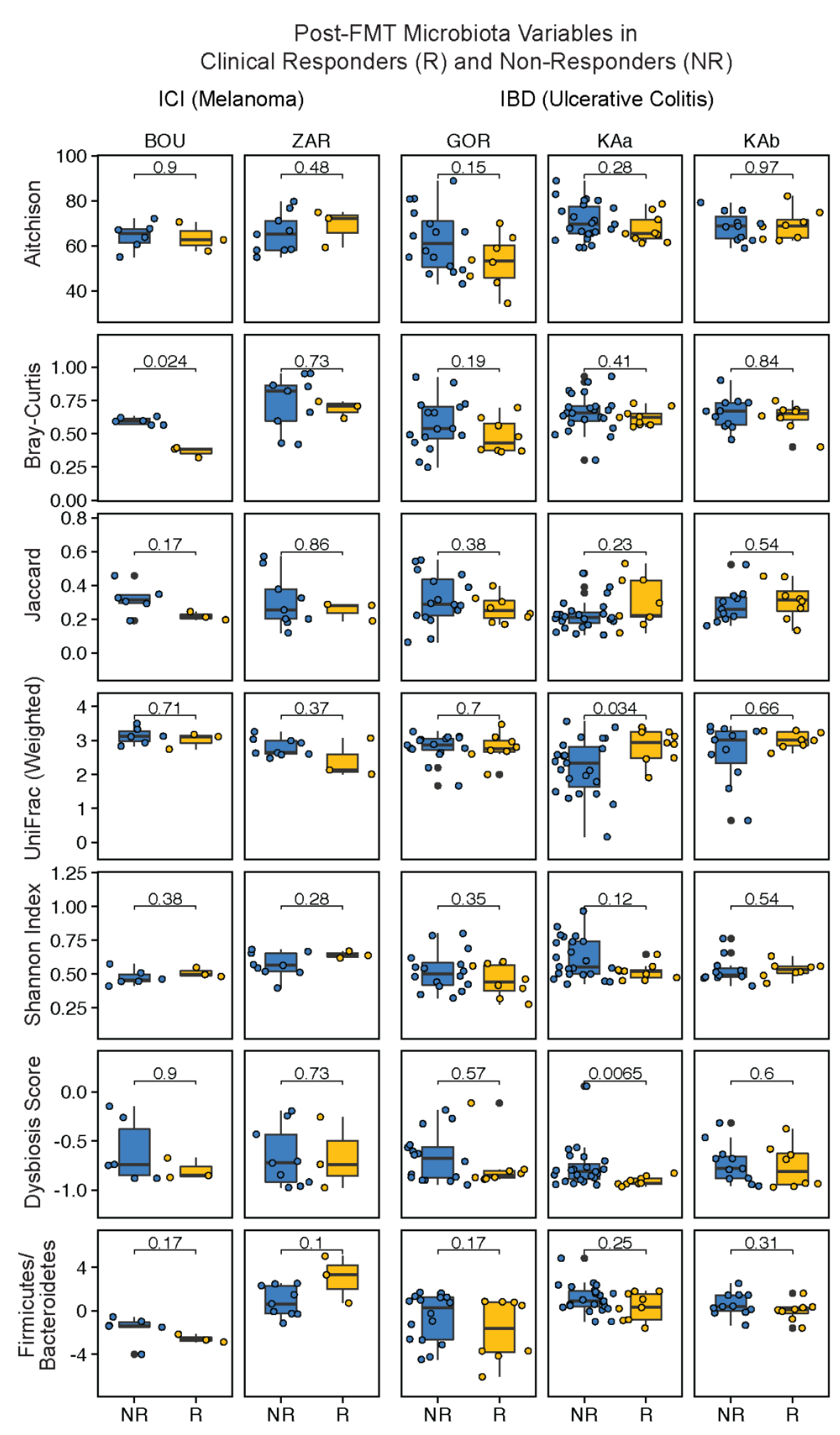
Comparison of taxonomic microbiota compositions between responders (R) and non-responders (NR) in ICI and IBD trials. α-diversity (Shannon Index), Dysbiosis Score, the *Firmicutes*/*Bacteroidetes* ratio, and β-diversity (Aitchison distance, Bray-Curtis dissimilarity, Jaccard distance, weighted UniFrac) between patients and their donors for Responder/Non-Responder patient groups (see Methods) within the ICI and IBD studies.

